# Does the choice of radiopharmaceutical affect the rate of incidental diagnosis of cardiac amyloidosis by bone scintigraphy?

**DOI:** 10.1101/2025.10.30.25338330

**Authors:** Andre Poon, Will Styles, Katherine Sneddon, Terence Doyle, Rathan M. Subramaniam, Sean Coffey

**Affiliations:** Department of Radiology, Dunedin Public Hospital, Health New Zealand Southern, New Zealand; Department of Medicine - HeartOtago, Dunedin School of Medicine, University of Otago, New Zealand; Department of Medicine, Dunedin School of Medicine, University of Otago, New Zealand; Department of Medicine – HeartOtago, Dunedin School of Medicine, University of Otago and Department of Cardiology, Dunedin Public Hospital, Health New Zealand Southern, New Zealand

## Abstract

**Background:** Cardiac radiopharmaceutical uptake during bone scintigraphy is a specific finding of cardiac amyloidosis, particularly transthyretin amyloid. There is little evidence to guide the optimum choice of technetium labelled radiopharmaceutical in protocols to detect cardiac amyloid. In our practice, different radiopharmaceuticals were used based on external factors such as availability and pricing. We wished to use this natural experiment to examine the rate of incidental cardiac uptake in clinically indicated bone scintigraphy.

**Methods:** Bone scintigraphy for non-cardiac indications was examined over a three-year period. Radiopharmaceutical and demographic data were recorded, and scans were evaluated retrospectively using the Perugini scale.

**Results:** One thousand four hundred seventy-three scans were examined, and of these, the radiopharmaceutical used was: 209 DPD, 175 HDP, 51 HMDP, and 1038 MDP. There were no significant differences between radiopharmaceutical groups regarding patient demographics or indications for imaging.

The rate of radiopharmaceutical cardiac uptake (Perugini grade >1) was 2.4% for DPD, 0% HDP, 0% HMDP, and 0.3% MDP (Fisher exact test p=0.011).

**Conclusions:** DPD may confer higher sensitivity than other radiopharmaceuticals to diagnose cardiac amyloid. These results encourage a direct comparison of radiopharmaceuticals to detect cardiac amyloid.

## Background

Cardiac transthyretin amyloidosis (ATTR-CA) has been increasingly recognised as a cause of significant morbidity and mortality.^1^ With the advent of new treatments that can alter the disease course, detection of ATTR-CA,^2^ whether suspected as the primary cause for a patient’s symptoms or detected incidentally, has entered a new era of importance.

Bone scintigraphy has an established role in diagnosing cardiac transthyretin amyloidosis, particularly when there is suspicion of the condition already.^3^ However, even incidental detection of ATTR-CA on bone scintigraphy has prognostic implications.^4^ Despite this, the optimal protocol for diagnosis of ATTR-CA has yet to be determined,^5,6^ in particular, the choice of the radiopharmaceutical. The 2019 ASNC/AHA/ASE/EANM/HFSA/ISA/SCMR/SNMMI Expert Consensus Recommendation for multimodality imaging in Cardiac Amyloidosis noted that “[a]lthough there is no direct comparison between these tracers, the information available suggests they can be used interchangeably” .^7^

In our practice, various radiopharmaceuticals (^99m^TcDPD, ^99m^TcHDP, ^99m^TcHMDP, and ^99m^TcMDP) were used based on patient-external factors, in particular availability and pricing. We wished to use this natural experiment to examine the rate of cardiac uptake in clinically indicated bone scans. We hypothesised that, although the population being imaged was similar, the use of DPD would result in a higher number of incidental diagnoses of ATTR-CA.

## Methods

### Patient selection

The study was a retrospective single centre study consisting of patients older than 40 years who underwent bone scintigraphy including the entire cardiac field for any indication by any referrer between 1 Sept 2017 to 31 Dec 2020 at Dunedin Public Hospital in New Zealand. We excluded scans where the radiopharmaceutical was not recorded or scans where cardiac amyloid was clinically suspected (i.e., this was the scan indication). The first scan was chosen for analysis in individuals undergoing repeated imaging for clinical reasons (most commonly, for follow-up of metastatic bone disease).

### Bone Scintigraphy

All bone scintigrams were performed using local protocols. Anterior and posterior whole-body planar views were obtained post 750-800 MBq intravenous injection of radiopharmaceutical after 2 hours.

In our nuclear medicine department, DPD, HDP, HMDP, and MDP radiopharmaceutical is procured in advance by the lead nuclear medicine technologist every six weeks and selected by availability and the lowest market price at the time. The radiopharmaceutical used in a given study was thereby allocated by the market cost of the radiopharmaceutical at the most recent procurement, rather than any patient or indication-related factors. As such, this allowed a natural experiment to be examined, where we assume a relatively stable cohort of patients and indications.

All scans were performed on GE Discovery MN/CT 670 Pro (GE Healthcare, Waukesha, Wi, USA).

### Visual score

Cardiac uptake of all bone scintigrams were graded using the Perugini grading scale (0 = no uptake, 1 = less than bone, 2 = equal to bone, 3 = greater than bone).^8^ A scan was identified as positive for cardiac uptake if the Perugini score was two or greater. Two investigators (AP and WS) graded all scans. A selection of 11 scans were cross-read by two consultant radiologists blinded to the original grading. Interrater reliability was good, with Fleiss’ kappa 0.79 for the three independent raters.

### Ethical approval

Ethics approval was obtained from the University of Otago Human Research Ethics Committee for this study (approval number: H21/084), which waived a requirement for individual participant consent.

### Statistical analyses

As this is a descriptive study, results were summarised using mean and standard deviation for age and percentages for categorical variables. We used one-way analysis of variation to test the null hypothesis of no difference in ages across the different radiopharmaceutical groups. We used the Fisher exact test to test the null hypothesis of no difference in gender, major indications for scanning, and rate of radiopharmaceutical uptake across the different radiopharmaceutical groups. A threshold of p<0.05 was chosen to represent statistical significance. Analyses were completed using R version 4.1.2.^9^

## Results

### Cohort

A total of 1689 bone scans with complete cardiac visualisation and radiopharmaceutical details were performed. Four scans were excluded as ATTR-CA was the clinical indication. After the exclusion of repeat scans, 1473 scans remained for analysis.

Results are shown in Table 1. The patients had a mean age of 68 years (standard deviation 13 years). 50.8% (748) of the scans were performed on women. According to the radiopharmaceutical group, there were no statistically significant differences in age or proportion of women.

**Table 1.**
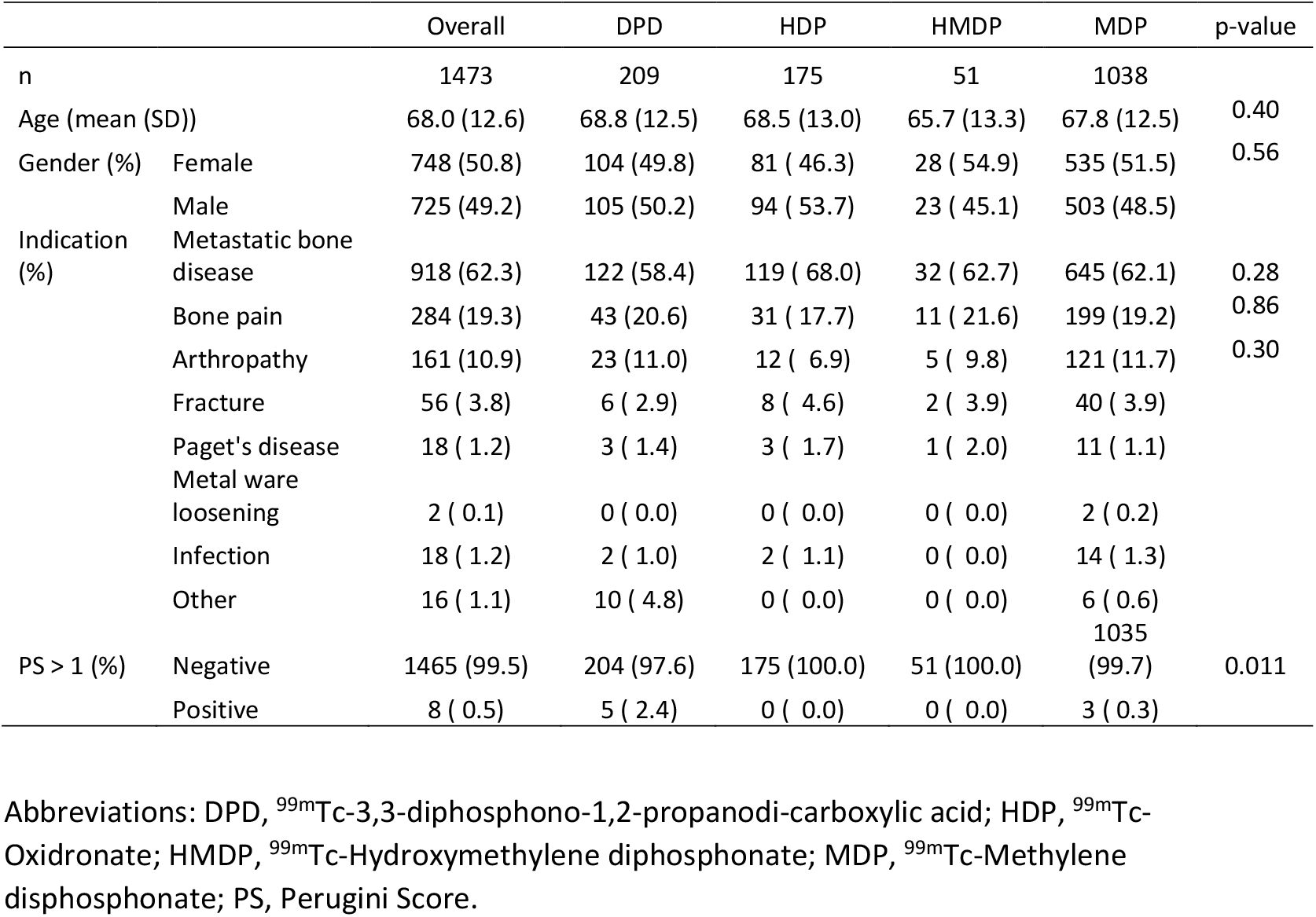
Demographics and cardiac uptake by radiopharmaceutical group.

The most common indications for imaging were identifying metastatic bone disease (62%, n=918); bone pain (19%, n=284); joint pain/arthropathy (11%, n=161); suspected fracture (4%, n=56); Paget’s disease (1%, n=18) and suspected infection (1%, n=18). There were no statistically significant differences in proportions of major indications (metastatic bone disease, bone pain, or arthropathy) between radiopharmaceutical groups.

### Radiopharmaceutical uptake

Overall, 8 patients (0.5%) had positive (Perugini grade 2 or 3) cardiac radiotracer uptake (Table 1). This was more common in studies using DPD with 5 (2.4%) positive compared to 0 positive in studies using HDP and HMDP and 3 (0.3%) in studies using MDP. This difference between groups was statistically significant (p=0.011).

## Discussion

In summary, we retrospectively examined clinically indicated bone scintigraphy performed for non-cardiac indications according to the use of radiopharmaceuticals. As our radiopharmaceutical choice was based on factors extrinsic to the patient, we were able to examine the results of a natural experiment. Despite similar patient populations, we found that there is a higher rate of cardiac radiotracer uptake using DPD compared to HMDP, PYP, or MDP.

Although not a formal randomised controlled trial, the effectively random assignment of radiotracer to individual scans provides scientific robustness to our findings. In particular, our results disagree with a 2021 meta-analysis that suggested that HMDP was more accurate for diagnosing cardiac amyloid.^5^ This suggestion was based on pooling studies performed in different patient cohorts, so confounding may well have entered the analysis. For example, the study that provided evidence of superiority of HMDP was rated as at risk of bias due to patient selection. In addition, participants had a median age of 75 years,^10^ compared to 64, 62, and 64 years for the three studies examined for diagnostic accuracy of DPD.^11–13^

## Limitations

The major limitation to the study is the relatively small number of patients with cardiac radiotracer uptake. Our observed rate is less than that seen in a study of older patients aged 70 years and older using HMDP (3.5%),^4^ but similar to that in a group with a more similar age range to ours (0.4%), imaged using DPD.^14^

## New Knowledge Gained

DPD may confer a higher sensitivity than HMDP, HDP, and MDP for the diagnosis of incidental ATTR-CA on bone scintigraphy.

## Conclusions

The choice of radiopharmaceutical may significantly affect cardiac radiotracer uptake and should be addressed in any future protocol. This study supports the need for a direct comparison of radiopharmaceuticals to detect cardiac amyloid.

## Data Availability

Data is not available due to New Zealand National Ethical Standards and relevant privacy legislation.

## Abbreviations

ATTR-CA: Cardiac transthyretin amyloidosis
^99m^Tc: ^99m^Technetium
DPD: ^99m^Tc-3,3-diphosphono-1,2-propanodi-carboxylic acid
HDP: ^99m^Tc-Oxidronate
HMDP: ^99m^Tc-Hydroxymethylene diphosphonate
MDP: ^99m^Tc-Methylene disphosphonate
CT: Computer Tomography
SPECT: Single-photon emission tomography

## Notes

Conflicts of interest: No authors have a conflict of interest to report

### Competing Interest Statement

The authors have declared no competing interest.

### Funding Statement

This study did not receive any funding.

### Author Declarations

The Human Research Ethics Committee of the University of Otago gave ethical approval for this work.

## References

1 Stassen J, van der Bijl P, Bax JJ. Optimizing 99mTc-DPD scintigraphy: Adding value to the diagnosis and treatment of cardiac transthyretin amyloidosis. J Nucl Cardiol 2021;28:2497–9. doi:10.1007/s12350-021-02716-5

2 Maurer MS, Schwartz JH, Gundapaneni B, et al. Tafamidis Treatment for Patients with Transthyretin Amyloid Cardiomyopathy. N Engl J Med 2018;379:1007–16. doi:10.1056/NEJMoa1805689

3 Dorbala S, Ando Y, Bokhari S, et al. ASNC/AHA/ASE/EANM/HFSA/ISA/SCMR/SNMMI expert consensus recommendations for multimodality imaging in cardiac amyloidosis: Part 2 of 2—Diagnostic criteria and appropriate utilization. J Nucl Cardiol 2020;27:659–73. doi:10.1007/s12350-019-01761-5

4 Suomalainen O, Pilv J, Loimaala A, et al. Prognostic significance of incidental suspected transthyretin amyloidosis on routine bone scintigraphy. J Nucl Cardiol Published Online First: 22 October 2020. doi:10.1007/s12350-020-02396-7

5 Zhao H, Hu H, Cui W. Performance of bone tracer for diagnosis and differentiation of transthyretin cardiac amyloidosis: a systematic review and meta-analysis. Diagnostic Interv Radiol 2021;27:802–10. doi:10.5152/dir.2021.20662

6 Schatka I, Bingel A, Schau F, et al. An optimized imaging protocol for [99mTc]Tc-DPD scintigraphy and SPECT/CT quantification in cardiac transthyretin (ATTR) amyloidosis. J Nucl Cardiol 2021;28:2483–96. doi:10.1007/s12350-021-02715-6

7 Dorbala S, Ando Y, Bokhari S, et al. ASNC/AHA/ASE/EANM/HFSA/ISA/SCMR/SNMMI expert consensus recommendations for multimodality imaging in cardiac amyloidosis: Part 1 of 2—evidence base and standardized methods of imaging. J Nucl Cardiol 2019;26:2065–123. doi:10.1007/s12350-019-01760-6

8 Perugini E, Guidalotti PL, Salvi F, et al. Noninvasive etiologic diagnosis of cardiac amyloidosis using 99mTc-3,3-diphosphono-1,2-propanodicarboxylic acid scintigraphy. J Am Coll Cardiol 2005;46:1076–84. doi:10.1016/j.jacc.2005.05.073

9 R Core Team. R: A Language and Environment for Statistical Computing. 2020. https://www.r-project.org/

10 Gallini C, Tutino F, Martone R, et al. Semi-quantitative indices of cardiac uptake in patients with suspected cardiac amyloidosis undergoing 99mTc-HMDP scintigraphy. J Nucl Cardiol 2021;28:90–9. doi:10.1007/s12350-019-01643-w

11 Moore PT, Burrage MK, Mackenzie E, et al. The Utility of 99m Tc-DPD Scintigraphy in the Diagnosis of Cardiac Amyloidosis: An Australian Experience. Hear Lung Circ 2017;26:1183–90. doi:10.1016/j.hlc.2016.12.017

12 de Haro-del Moral FJ, Sánchez-Lajusticia A, Gómez-Bueno M, et al. Role of cardiac scintigraphy with ^99^mTc-DPD in the differentiation of cardiac amyloidosis subtype. Rev Esp Cardiol (Engl Ed) 2012;65:440–6. doi:10.1016/j.recesp.2011.12.015

13 Rapezzi C, Quarta CC, Guidalotti PL, et al. Usefulness and limitations of 99mTc-3,3-diphosphono-1,2-propanodicarboxylic acid scintigraphy in the aetiological diagnosis of amyloidotic cardiomyopathy. Eur J Nucl Med Mol Imaging 2011;38:470–8. doi:10.1007/s00259-010-1642-7

14 Longhi S, Guidalotti PL, Quarta CC, et al. Identification of TTR-related subclinical amyloidosis with 99mTc-DPD scintigraphy. JACC Cardiovasc Imaging 2014;7:531–2. doi:10.1016/j.jcmg.2014.03.004

